# Severity of tauopathy differs between logopenic variant primary progressive aphasia individuals with or without a history of learning differences

**DOI:** 10.64898/2026.03.03.26347280

**Authors:** Salvatore Spina, Zachary A. Miller, Sevinç Jakab, Margherita Tamagnini, Maria Luisa Mandelli, Lisa Kritikos, Hieu Pham, Siddarth Ramkrishnan, Mia Lin, Jaeyeon Kim, Mercedes Paredes, Howie J. Rosen, Lea T. Grinberg, William W. Seeley, Bruce L. Miller, Maria Luisa Gorno-Tempini

## Abstract

**OBJECTIVE:** To assess differences in the severity of Alzheimer’s disease neuropathological changes in disease epicenters of patients with logopenic variant PPA (lvPPA) with a history of learning differences/developmental dyslexia (LD) versus lvPPA patients without such history (non-LD).

**BACKGROUND:** Learning differences, including developmental dyslexia, are overrepresented in the lvPPA population. It is not known whether a history of developmental differences is associated with a more severe phenotypic expression of Alzheimer’s disease pathology.

**DESIGN/METHODS:** We quantified the cortical area fraction of phospho-tau immunohistochemistry (IHC) and beta-amyloid IHC in the angular gyrus and superior temporal gyrus of postmortem brains of 15 cases of lvPPA secondary to Alzheimer’s disease of which 9 non-LD cases (2 males and 7 females), and 6 LD cases (2 males and 4 females). Histological sections were digitally acquired, and foreground IHC-signal was automatically separated and thresholded to quantify the respective tau and beta-amyloid area fractions in each region.

**RESULTS:** There were no differences in the mean age at death between the two groups. Disease duration was longer in the LD group (10.7 ± 1.2 years) than in the non-LD group (8.1 ± 0.8 years), *p*=0.09. When corrected for sex, age at death and Apo E4 carrier status, the LD group showed higher tau pathology burden in the superior temporal gyrus compared to the non-LD group (0.91% ± 0.37, *p*=0.03). No differences in tau pathology burden between the groups were observed in the angular gyrus (0.39% ± 0.41, *p*=0.37). There were no statistically significant differences in the area fraction of beta-amyloid between the two groups of patients in both the angular gyrus and the superior temporal gyrus.

**CONCLUSIONS:** Our data suggest that patients with lvPPA secondary to Alzheimer’s disease and a history of developmental differences have higher tau-pathology burden in the superior temporal gyrus compared to lvPPA-AD patients without such history.

## INTRODUCTION

Logopenic variant primary progressive aphasia (lvPPA) is a neurodegenerative syndrome within the primary progressive aphasia spectrum, characterized by impaired single-word retrieval, phonological errors, and disproportionately impaired sentence and phrase repetition ^1^. Unlike the semantic and nonfluent/agrammatic variants, lvPPA is most commonly associated with Alzheimer’s disease (AD) pathology rather than frontotemporal lobar degeneration ^2–4^. Converging evidence from structural MRI, FDG-PET, and tau PET studies indicates that lvPPA reflects a focal AD phenotype defined by early tau accumulation and neurodegeneration in left temporoparietal regions that support phonological working memory, lexical access, and auditory–verbal short-term memory ^3, 5^. These observations underscore the importance of intrinsic network vulnerabilities in shaping regional patterns of AD-related tau deposition.

Within this framework of selective network vulnerability, a growing body of literature has explored whether early-life neurodevelopmental differences may influence which brain networks become most susceptible to neurodegenerative disease decades later^6–8^. Developmental dyslexia, a common learning disability characterized by deficits in phonological processing, grapheme– phoneme integration, and fluent reading, represents one such factor that may shape life-long organization of language networks ^9^. Neuroimaging studies of dyslexia consistently reveal atypical structural and functional profiles within left temporoparietal and occipitotemporal regions, including reduced gray matter volume, altered white-matter microstructure, and reduced activation during phonological tasks ^10–12^. Notably, these regions overlap extensively with those that show early degeneration in lvPPA, raising the possibility that long-standing neurodevelopmental variation might modulate the expression or severity of later-life AD pathology^7^.

Emerging epidemiologic and clinicopathologic observations further suggest a potential link between neurodevelopmental language differences and later-life aphasic neurodegenerative syndromes^13, 14^. A history of learning disability or developmental dyslexia has been reported at increased frequency in individuals with PPA compared to cognitively normal controls, raising the hypothesis that early-life language network organization may confer regionally selective vulnerability under proteinopathy stress^7, 15–17^. While direct longitudinal evidence connecting dyslexia to increased AD incidence remains limited, the anatomical convergence between dyslexia-related cortical differences and temporo-parietal regions preferentially affected in logopenic variant PPA—most commonly associated with AD pathology—provides a mechanistic framework for investigating developmental–degenerative interactions^1, 7, 15, 18^. Despite these intriguing observations, the relationship between dyslexia and the biological underpinnings of lvPPA— particularly tau pathology—remains largely unexamined.

Tau pathology is the principal substrate associated with symptom progression in AD and its clinical variants^19^. In lvPPA, tau deposition—as measured in vivo through tau PET or assessed post-mortem—is disproportionately concentrated in left posterior superior temporal, inferior parietal, and temporoparietal junction regions critical for phonological processing.^5^ In addition, unique patterns of tau deposition have been observed in individuals with history of developmental differences associated with cortical developmental abnormalities and/or a late-in-life history of lvPPA^14, 20, 21^. Nevertheless, whether a history of dyslexia meaningfully influences the magnitude or regional distribution of tau pathology in lvPPA has not yet been empirically tested.

This study aims to verify whether tau pathology differs between individuals with lvPPA who have a documented or reported history of dyslexia and those without such a history. Using quantitative measures of tau pathology, we examine both total burden and regional distribution across language-related cortical networks. By investigating these potential differences, this study seeks to illuminate how early-life neural architecture may intersect with AD pathobiology, ultimately contributing to more refined models of disease susceptibility and variability within lvPPA.

## METHODS

We searched the UCSF Neurodegenerative Disease Brain Bank for cases with a primary pathological diagnosis of Alzheimer’s disease and a clinical diagnosis of logopenic variant primary progressive aphasia.^1, 22–25^ Twenty-two brains were identified. Brains were obtained post-mortem and processed as previously described, from patients enrolled during life in longitudinal studies on dementia at the Fein Memory and Aging Center, UCSF, approved by the UCSF Institutional Review Board.^26^ Tau immunohistochemistry in both regions of interest, the angular gyrus and the superior temporal gyrus, as well as information on sex, age at death, disease duration, and Apo E genotype was available for only 15 of these 22 cases, which were therefore included in the study. Beta-amyloid immunohistochemistry was available for 12 of the 15 cases in the superior temporal gyrus and for 13 of the 15 cases in the angular gyrus. Details on the pathological assessment of the 15 cases in the study are provided in Table 1. Information regarding a history of developmental dyslexia or language-related learning disabilities was retrospectively obtained through review of clinical information as previously described^7, 8, 14^.

**Table 1.** Subjects’ demographics and neuropathological data. Group: 0 = non-learning differences), 1 = learning differences. Sex: M =male, F = female. NFT: neurofibrillary tangles. CERAD NP: Consortium to Establish a Registry for Alzheimer’s Disease, neuritic plaques. LBD: Lewy body disease. Amygdala: amygdala-only. LATE: limbic-predominant aging-related TDP43 encephalopathy. AGD: argyrophilic grain disease. IHC: immunohistochemistry. STG: superior temporal gyrus. AG: angular gyrus. −: absent. + : present, limbic type. n.a: not available. * : available.

| Subject | Group | Sex | Apo E4 carrier | Thal phase | Braak NFT stage | CERAD NP score | LBD stage | LATE | AGD | Tau-IHC |  | Beta-amyloid IHC |  |
| --- | --- | --- | --- | --- | --- | --- | --- | --- | --- | --- | --- | --- | --- |
|  |  |  |  |  |  |  |  |  |  | STG | AG | STG | AG |
| 1 | 0 | M | 0 | n.a. | 6 | 3 | 1 | - | - | * | * | n.a. | * |
| 2 | 1 | F | 1 | 5 | 6 | 3 | 0 | - | - | * | * | n.a. | * |
| 3 | 1 | F | 1 | 5 | 6 | 3 | 0 | - | - | * | * | * | * |
| 4 | 1 | M | 0 | 5 | 6 | 3 | 3 | - | - | * | * | * | * |
| 5 | 0 | F | 0 | 5 | 6 | 3 | 0 | - | + | * | * | * | * |
| 6 | 1 | F | 0 | 5 | 6 | 3 | Amygdala | - | - | * | * | * | * |
| 7 | 1 | M | 0 | 5 | 6 | 3 | 0 | - | + | * | * | * | * |
| 8 | 0 | F | 0 | 5 | 6 | 3 | 0 | - | + | * | * | * | * |
| 9 | 0 | F | 1 | 5 | 6 | 3 | 3 | - | + | * | * | * | * |
| 10 | 1 | F | 1 | 5 | 6 | 3 | 0 | - | + | * | * | * | * |
| 11 | 0 | F | 1 | 5 | 6 | 3 | 0 | - | + | * | * | * | * |
| 12 | 0 | F | 0 | 5 | 6 | 3 | 0 | - | + | * | * | * | * |
| 13 | 0 | F | 1 | 5 | 6 | 3 | Amygdala | - | - | * | * | n.a. | n.a. |
| 14 | 0 | M | 1 | 5 | 6 | 3 | 0 | - | + | * | * | * | n.a. |
| 15 | 0 | F | 0 | 4 | 6 | 3 | 0 | - | + | * | * | * | * |

Standard 8-micron archival histological slides of the inferior frontal gyrus, the angular gyrus and the superior temporal gyrus stained for tau-immunohistochemistry (CP13) or beta-amyloid immunohistochemistry (4G8) were digitally acquired using a Zeiss Axioscan 7 system (Carl Zeiss Microscopy GmbH, Jena, Germany) at 20X magnification. Regions of interests were then manually drawn to capture the cortical ribbon in all slides, with care to avoid regions of lack of perpendicularity of the section or gross artifacts and used for area fraction quantification of the IHC signal. Images were exported in tiles as .tiff format. For each staining and region and case, 10% of the tiles images were randomly selected to be included in the analyses. These tile images were run though a customized Python script for color deconvolution based on scikit-image library.^27, 28^ DAB channel images, which contain the phospho-tau staining, were thresholded with a customized Python script based on maximum entropy thresholding method (Figure 1).^29, 30^ Finally, the percentage area occupied (%AO) per tile was calculated based on [(number of IHC positive pixels / total number of pixels) x 100] and average of all the tiles was calculated for the final %AO per patient and brain region.

**Figure 1.**
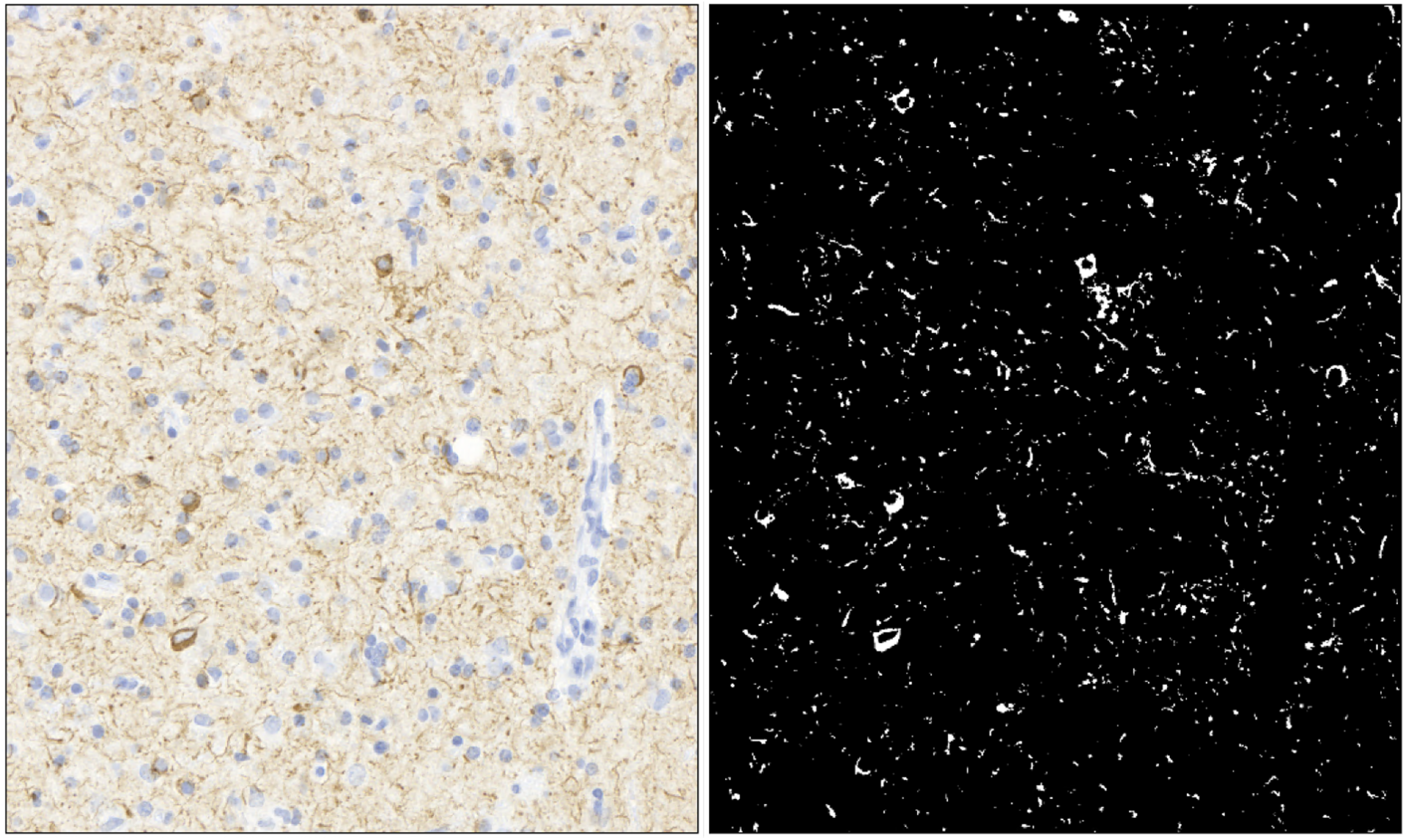
Left: tau-immunohistochemistry (CP13). Right: segmentation mask of tau immune-positive signal following color deconvolution, and DAB channel images thresholding using a customized Python script based on maximum entropy thresholding method.

Demographics differences were assessed by t-test analyses. Differences in the amount of tau pathology burden or beta-amyloid burden in the regions of interest between the two groups were assessed through liner regression, adjusting for sex, age at death, Apo E status and disease duration.

## RESULTS

### Subjects’ demographics

A history of developmental dyslexia or learning differences (LD-group)) was retrospectively confirmed in 6 patients (2 males and 4 females) and ruled out in the remaining 9 (2 males and 7 females) of these patients (non-LD group) through review of their clinical information. There was no statistically significant difference in the age at death between the two groups (LD = 67.3 ± 2.5 years; non-LD group = 67.6 ± 2.4 years, *p* = 0.95). The disease duration was longer in the LD group (10.7 ± 1.2 years) than in the non-LD group (8.1 ± 0.8 years, *p* = 0.09), although the difference did not reach statistical significance. There were 4 Apo E 4 carriers in the non-LD group (44.4%), and 3 in the LD group (50%).

### Tau pathology burden is higher in the superior temporal gyrus of lvPPA patients with a history of developmental differences compared to those without such history

When adjusting for sex, age at death, and Apo E status lvPPA patients with a history of LD have a statistically significant higher tau pathological burden in the superior temporal gyrus compared to lvPPA patients without such history (n=15, estimate = 0.91 ± 0.37, CI = 0.09 – 1.73, *p* = 0.03). Violin plots are depicted in Figure 2. A higher amount of tau pathology was found in females compared to males (n= 15, estimate = 0.50 ± 0.42, CI = −0.44 – 1.44, *p* = 0.27), and in Apo E4 carriers compared to non-carriers (n= 15, estimate = 0.20 ± 0.38, CI = −0.63 – 1.04, *p* = 0.60) although these differences did not reach statistical significance. There was a trend to a lower tau pathological burden with increasing age at death (n= 15, estimate = −0.05 ± 0.03, CI = −0.11 – 0.02, *p* = 0.13).

**Figure 2.**
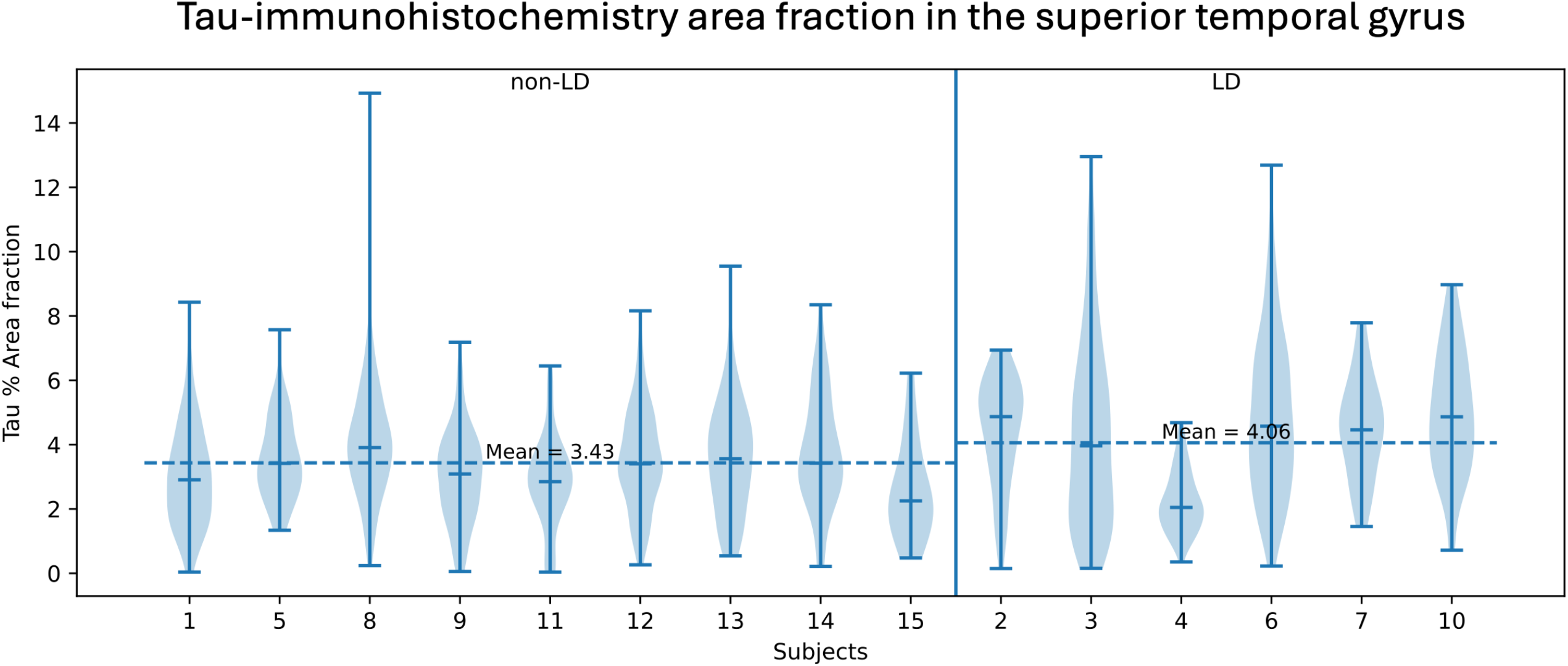
Violin-plot depicting the distribution of tau area fraction in the superior temporal gyrus samples across the individual tile images selected post randomization for each of the participants in the study.

When adding disease duration as a covariate to the above regression model, a trend for a higher tau pathological burden in the superior temporal gyrus of lvPPA patients with a history of LD compared to those without such history was also observed (n= 15, estimate = 0.65 ± 0.41, CI = −0.27 – 1.57, *p* = 0.15). A higher amount of tau pathology was found in females compared to males (n= 15, estimate = 0.37 ± 0.42, CI = −0.59 – 1.32, *p* = 0.41), and in Apo E4 carriers compared to non-carriers (n= 15, estimate = 0.24 ± 0.36, CI = −0.59 – 1.06, *p* = 0.53), together with a trend toward higher tau pathology burden with increased disease duration (n= 15, estimate = 0.10 ± 0.07, CI = −0.07 – 0.26, *p* = 0.23) although these differences did not reach statistical significance. There was a trend to a lower tau pathological burden with increasing age at death (n= 15, estimate = −0.04 ± 0.03, CI = −0.10 – 0.02, *p* = 0.19).

### lvPPA patients with or without a history of developmental differences have no different severity of tau pathology burden in the angular gyrus

When comparing the amount of tau pathology burden in the angular gyrus of lvPPA patients with a history of developmental differences to those without such history, adjusting for sex, age at death, and Apo E status, no statistically significant differences were observed (n= 15, estimate = 0.39 ± 0.41, CI = −0.52 – 1.31, *p* = 0.37). Violin plots are depicted in Figure 3. A higher amount of tau pathology was found in females compared to males (n= 15, estimate = 0.40 ± 0.47, CI = −0.65 – 1.45, *p* = 0.42), while a lower amount of tau pathology was associated with an Apo E4 carrier status compared to non-carriers (n= 15, estimate = −0.01 ± 0.42, CI = −0.94 – 0.93, *p* = 0.98) and increasing age (n= 15, estimate = −0.02 ± 0.03, CI = −0.09 – 0.05, *p* = 0.51), although none of these differences reached statistical significance.

**Figure 3.**
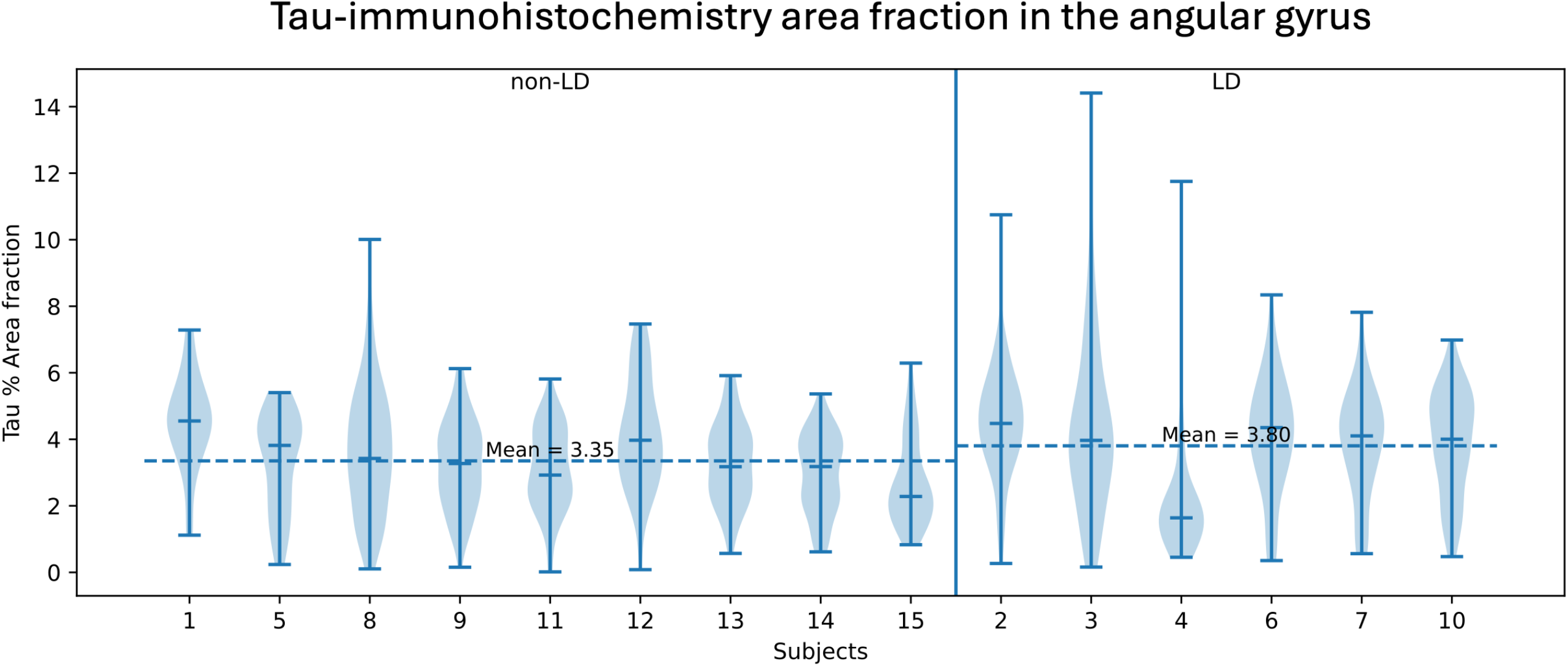
Violin-plot depicting the distribution of tau area fraction in the angular gyrus samples across the individual tile images selected post randomization for each of the participants in the study.

When adding disease duration as a covariate to the above regression model, we found again no statistically significant differences in the amount of tau pathology burden between the two groups (n= 15, estimate = −0.11 ± 0.36, CI = −0.92 – 0.70, *p* = 0.77). Higher disease duration was associated with a higher tau pathology burden (n= 15, estimate = 0.18 ± 0.06, CI = 0.04 – 0.33, *p* = 0.02). No statistically significant differences were associated with sex (n= 15, estimate = 0.15 ± 0.37, CI = −0.69 – 0.99, *p* = 0.70), age at death (n= 15, estimate = −0.01 ± 0.02, CI = −0.06 – 0.05, *p* = 0.77), or Apo E carrier status (n= 15, estimate = 0.05 ± 0.32, CI = −0.67 – 0.78, *p* = 0.88).

### lvPPA patients with or without a history of developmental differences have no different severity of beta-amyloid pathology burden in the superior temporal gyrus

When comparing the amount of beta-amyloid pathology burden in the superior temporal gyrus of lvPPA patients with a history of developmental differences to those without such history, adjusting for sex, age at death, and Apo E status, no statistically significant differences were observed (n= 12, estimate = −1.67 ± 1.54, CI = −5.32 – 1.99, *p* = 0.32). A lower amount of beta-amyloid pathology was found in females compared to males (n= 12, estimate = −0.50 ± 1.77, CI = −4.68 – 3.68, *p* = 0.79), Apo E4 carriers (n= 12, estimate = −0.09 ± 1.48, CI = −3.60 – 3.42, *p* = 0.96), and increasing age (n= 12, estimate = −0.09 ± 0.12, CI = −0.37 – 0.19, *p* = 0.46), although none of these differences reached statistical significance.

When adding disease duration as a covariate to the above regression model, we found again no statistically significant differences in the amount of beta-amyloid pathology burden between the two groups (n= 12, estimate = −1.09 ± 2.27, CI = −6.65 – 4.46, *p* = 0.65). No statistically significant differences were associated with sex (n= 12, estimate = 0.05 ± 2.41, CI = −5.85 – 5.95, *p* = 0.98), age at death (n= 12, estimate = −0.11 ± 0.14, CI = −0.45 – 0.22, p = 0.44), Apo E carrier status (n= 12, estimate = −0.01 ± 1.60, CI = −3.92 – 3.90, *p* = 0.99), and disease duration (n= 12, estimate = −0.15 ± 0.43, CI = −1.21 – 0.89, p = 0.21).

### lvPPA patients with or without a history of developmental differences have no different severity of beta-amyloid pathology burden in the angular gyrus

When comparing the amount of beta-amyloid pathology burden in the angular gyrus of lvPPA patients with a history of developmental differences to those without such history, adjusting for sex, age at death, and Apo E status, no statistically significant differences were observed (n= 13, estimate = −0.17 ± 1.05, CI = −2.59 – 2.25, *p* = 0.88). No statistically significant differences were associated with sex (n= 13, estimate = 1.78 ± 1.35, CI = −1.32 – 4.90, *p* = 0.22), age at death (n= 13, estimate = −0.08 ± 0.73, CI = −0.25 – 0.85, *p* = 0.29), and Apo E carrier status (n= 13, estimate = 0.20 ± 1.17, CI = −2.49 – 2.90, *p* = 0.87).

When adding disease duration as a covariate to the above regression model, we found again no statistically significant differences in the amount of beta-amyloid pathology burden between the two groups (n= 13, estimate = 0.28 ± 1.17, CI = −2.74 – 2.80, *p* = 0.98). No statistically significant differences were associated with sex (n= 13, estimate = 1.84 ± 1.42, CI = −1.52 – 5.20, *p* = 0.24), age at death (n= 13, estimate = −0.09 ± 0.08, CI = −0.27 – 0.09, *p* = 0.28), Apo E carrier status (n= 13, estimate = 0.28 ± 1.23, CI = −2.65 – 3.21, *p* = 0.83), and disease duration (n= 13, estimate = −0.10 ± 0.21, CI = −0.59 – 0.39, *p* = 0.63).

## Discussion

In this study, we demonstrate that lvPPA cases with a documented history of learning differences including developmental dyslexia exhibit a significantly higher burden of tau aggregates in the superior temporal gyrus (STG) compared to lvPPA cases without such a history. These findings provide novel neuropathological evidence supporting the hypothesis that neurodevelopmental language disorders may confer selective vulnerability to later-in-life neurodegenerative processes, particularly within language-related cortical networks.

The STG is a critical hub for phonological processing and auditory–verbal integration, functions consistently implicated in both developmental dyslexia and lvPPA ^1, 31^. Neuroimaging studies of dyslexia have repeatedly demonstrated structural and functional abnormalities within the left STG and surrounding temporoparietal cortex, including altered cortical thickness, atypical activation during phonological tasks, and disrupted connectivity ^11, 32, 33^. Our findings extend this literature by suggesting that early-life alterations in this region may predispose the same cortical territory to increased tau aggregation in the context of Alzheimer’s disease–related pathology underlying lvPPA ^18, 34^.

These results are consistent with network-based models of neurodegeneration, which posit that pathological proteins accumulate preferentially within large-scale brain networks that are metabolically active, highly interconnected, or developmentally vulnerable ^35, 36^. The language network targeted in lvPPA overlaps substantially with regions implicated in dyslexia, raising the possibility that longstanding network inefficiencies or compensatory hyperactivity may increase susceptibility to tau-related neurotoxicity later in life^10, 14^.

Several mechanisms may account for the increased tau burden observed in the STG of dyslexia-associated lvPPA cases. First, activity-dependent tau release and trans-synaptic propagation have been demonstrated in experimental models, suggesting that lifetime patterns of neuronal activity could influence regional tau accumulation ^37, 38^.

Developmental dyslexia has a significant heritable component, and among the most replicated susceptibility loci are DCDC2 and KIAA0319, both implicated in aspects of neuronal differentiation, neurite outgrowth, and structural connectivity during early brain development. Variants at these loci have been associated with differences in white matter microstructure and reading-related neural pathways, suggesting effects on neuronal development and circuit organization.^39^

Converging evidence from neurodegenerative disease research shows that cellular proteostasis mechanisms — including chaperone systems, protein clearance pathways, and axonal transport dynamics — critically determine neuronal resilience to misfolded proteins and pathological tau aggregation.^40^ Although no direct study has yet linked dyslexia-associated alleles to altered proteostatic function or tau homeostasis, the convergence of dyslexia gene functions on microtubule dynamics, neurite integrity, and cortical network connectivity raises a testable hypothesis: that subtle, developmentally mediated differences in neuronal architecture and transport capacity reduce the ability of affected neurons to cope with proteostatic stress, thereby increasing susceptibility to tau hyperphosphorylation and aggregation under later-life stress conditions^10, 14^. Importantly, the regional specificity of our findings argues against a generalized increase in tau pathology among lvPPA cases with dyslexia. Instead, the preferential involvement of the STG supports a model in which neurodevelopmental differences influence the topography of tau deposition rather than its overall burden. Developmental history may therefore act as a modifier of regional tau expression within an otherwise shared molecular disease process^14^.

Our findings also provide neuropathological support for clinical observations linking developmental learning disorders with primary progressive aphasia. Several studies have reported an increased prevalence of dyslexia or learning disabilities among patients with PPA, particularly the logopenic variant ^6, 7, 15^. However, pathological correlates of this association have remained largely unexplored. By demonstrating increased tau aggregation in a core phonological processing region, our data offer a potential biological substrate for these epidemiological and clinical associations.

This study has several limitations. Dyslexia history was ascertained retrospectively through clinical records and informant report, which may lead to underdiagnosis or misclassification, particularly in older cohorts educated prior to widespread recognition of dyslexia. Additionally, while tau burden was quantified postmortem, we cannot determine whether increased aggregation reflects earlier disease onset, accelerated accumulation, or impaired clearance. Longitudinal studies incorporating tau PET imaging in at-risk populations may help clarify the temporal dynamics of these processes ^41^. Finally, our sample size limits exploration of interactions with sex, education, or APOE genotype, all of which may influence tau distribution and clinical phenotype.

In conclusion, this study demonstrates increased tau aggregation in the superior temporal gyrus of lvPPA cases with a history of developmental dyslexia, supporting the concept that neurodevelopmental language vulnerabilities can shape the anatomical distribution of tau pathology decades later. These findings emphasize the importance of considering lifelong brain organization and developmental history when interpreting selective patterns of neurodegeneration and suggest that early-life cognitive traits may meaningfully modify disease expression in lvPPA.

## Data Availability

All data produced in the present study are available upon reasonable request to the authors

http://memory.ucsf.edu/resources/data

## Acknowledgments

This work was supported by the National Institutes of Health (NIH): R01NS050915 and P30AG062422. We are grateful to our patients for participating in this research. Samples from the National Centralized Repository for Alzheimer’s Disease and Related Dementias (NCRAD), which receives government support under a cooperative agreement grant (U24AG21886) awarded by the NIA, were used in this study.

## Conflict of interest statement

The authors have no conflict of interest related to this publication.

## Data Sharing Statement

UCSF Fein MAC ethics approval does not permit depositing anonymized study data in a public repository. Requests for access can be submitted through the UCSF-MAC Resource Portal (Request form: http://memory.ucsf.edu/resources/data). Access will be granted following UCSF-regulated procedures in accordance with ethical guidelines for the reuse of sensitive data. Researchers seeking access must submit a Material Transfer Agreement, which is available at: https://icd.ucsf.edu/material-transfer-and-data-agreements.

## Notes

### Competing Interest Statement

The authors have declared no competing interest.

### Funding Statement

This work was supported by the National Institutes of Health (NIH): R01NS050915 and P30AG062422. Samples from the National Centralized Repository for Alzheimer's Disease and Related Dementias (NCRAD), which receives government support under a cooperative agreement grant (U24AG21886) awarded by the NIA, were used in this study.

### Author Declarations

The Institutional Review Board (IRB) of University of California, San Francisco gave ethical approval for this work

## References

1. Gorno-Tempini ML, Hillis AE, Weintraub S, et al. Classification of primary progressive aphasia and its variants. Neurology 2011;76:1006–1014.

2. Mesulam MM, Rogalski EJ, Wieneke C, et al. Primary progressive aphasia and the evolving neurology of the language network. Nat Rev Neurol 2014;10:554–569.

3. Spinelli EG, Mandelli ML, Miller ZA, et al. Typical and atypical pathology in primary progressive aphasia variants. Ann Neurol 2017;81:430–443.

4. Rogalski E, Sridhar J, Rader B, et al. Aphasic variant of Alzheimer disease: Clinical, anatomic, and genetic features. Neurology 2016;87:1337–1343.

5. Ossenkoppele R, Cohn-Sheehy BI, La Joie R, et al. Atrophy patterns in early clinical stages across distinct phenotypes of Alzheimer’s disease. Hum Brain Mapp 2015;36:4421–4437.

6. Miller ZA, Ossenkoppele R, Graff-Radford NR, et al. Neurodevelopment and neural environment inform Alzheimer’s disease age at onset and phenotype. Alzheimers Dement 2025;21:e70668.

7. Miller ZA, Mandelli ML, Rankin KP, et al. Handedness and language learning disability differentially distribute in progressive aphasia variants. Brain 2013;136:3461–3473.

8. Miller ZA, Rosenberg L, Santos-Santos MA, et al. Prevalence of Mathematical and Visuospatial Learning Disabilities in Patients With Posterior Cortical Atrophy. JAMA Neurol 2018;75:728–737.

9. Richlan F. Developmental dyslexia: dysfunction of a left hemisphere reading network. Front Hum Neurosci 2012;6.

10. Paulesu E, Démonet JF, Fazio F, et al. Dyslexia:: Cultural diversity and biological unity. Science 2001;291:2165–2167.

11. Vandermosten M, Boets B, Wouters J, Ghesquiere P. A qualitative and quantitative review of diffusion tensor imaging studies in reading and dyslexia. Neurosci Biobehav Rev 2012;36:1532–1552.

12. Miller ZA, Hinkley LBN, Borghesani V, et al. Non-right-handedness, male sex, and regional, network-specific, ventral occipito-temporal anomalous lateralization in adults with a history of reading disability. Cortex 2025;183:116–130.

13. Mesulam MM, Weintraub S. Spectrum of primary progressive aphasia. Baillieres Clin Neurol 1992;1:583–609.

14. Miller ZA, Spina S, Pakvasa M, et al. Cortical developmental abnormalities in logopenic variant primary progressive aphasia with dyslexia. Brain Commun 2019;1.

15. Rogalski E, Johnson N, Weintraub S, Mesulam M. Increased frequency of learning disability in patients with primary progressive aphasia and their first-degree relatives. Arch Neurol 2008;65:244–248.

16. Miller ZA, Rankin KP, Graff-Radford NR, et al. TDP-43 frontotemporal lobar degeneration and autoimmune disease. J Neurol Neurosur Ps 2013;84:956–962.

17. Mesulam MM. Primary progressive aphasia--a language-based dementia. N Engl J Med 2003;349:1535–1542.

18. Mesulam M, Wicklund A, Johnson N, et al. Alzheimer and frontotemporal pathology in subsets of primary progressive aphasia. Ann Neurol 2008;63:709–719.

19. Petersen C, Nolan AL, de Paula Franca Resende E, et al. Alzheimer’s disease clinical variants show distinct regional patterns of neurofibrillary tangle accumulation. Acta Neuropathol 2019;138:597–612.

20. Sen A, Thom M, Martinian L, et al. Pathological tau tangles localize to focal cortical dysplasia in older patients. Epilepsia 2007;48:1447–1454.

21. Iyer A, Prabowo A, Anink J, Spliet WG, van Rijen PC, Aronica E. Cell injury and premature neurodegeneration in focal malformations of cortical development. Brain Pathol 2014;24:1–17.

22. Montine TJ, Phelps CH, Beach TG, et al. National Institute on Aging-Alzheimer’s Association guidelines for the neuropathologic assessment of Alzheimer’s disease: a practical approach. Acta Neuropathol 2012;123:1–11.

23. Braak H, Alafuzoff I, Arzberger T, Kretzschmar H, Del Tredici K. Staging of Alzheimer disease-associated neurofibrillary pathology using paraffin sections and immunocytochemistry. Acta Neuropathol 2006;112:389–404.

24. Thal DR, Rub U, Orantes M, Braak H. Phases of A beta-deposition in the human brain and its relevance for the development of AD. Neurology 2002;58:1791–1800.

25. Mirra SS, Heyman A, McKeel D, et al. The Consortium to Establish a Registry for Alzheimer’s Disease (CERAD). Part II. Standardization of the neuropathologic assessment of Alzheimer’s disease. Neurology 1991;41:479–486.

26. Spina S, La Joie R, Petersen C, et al. Comorbid neuropathological diagnoses in early versus late-onset Alzheimer’s disease. Brain 2021;144:2186–2198.

27. Ruifrok AC, Johnston DA. Ouantification of histochemical staining by color deconvolution. Anal Ouant Cytol Histol 2001;23:291–299.

28. van der Walt S, Schonberger JL, Nunez-Iglesias J, et al. scikit-image: image processing in Python. Peerj 2014;2.

29. Yaniv Z, Lowekamp BC, Johnson HJ, Beare R. SimpleITK Image-Analysis Notebooks: a Collaborative Environment for Education and Reproducible Research. J Digit Imaging 2018;31:290–303.

30. Lowekamp BC, Chen DT, Ibáñez L, Blezek D. The Design of SimpleITK. Front Neuroinform 2013;7.

31. Hickok G, Poeppel D. The cortical organization of speech processing. Nat Rev Neurosci 2007;8:393–402.

32. Shaywitz BA, Shaywitz SE, Pugh KR, et al. Disruption of posterior brain systems for reading in children with developmental dyslexia. Biol Psychiat 2002;52:101–110.

33. Richlan F. The Functional Neuroanatomy of Developmental Dyslexia Across Languages and Writing Systems. Front Psychol 2020;11:155.

34. Giannini LAA, Irwin DJ, McMillan CT, et al. Clinical marker for Alzheimer disease pathology in logopenic primary progressive aphasia. Neurology 2017;88:2276–2284.

35. Seeley WW, Crawford RK, Zhou J, Miller BL, Greicius MD. Neurodegenerative diseases target large-scale human brain networks. Neuron 2009;62:42–52.

36. Raj A, Kuceyeski A, Weiner M. A network diffusion model of disease progression in dementia. Neuron 2012;73:1204–1215.

37. Pooler AM, Phillips EC, Lau DH, Noble W, Hanger DP. Physiological release of endogenous tau is stimulated by neuronal activity. EMBO Rep 2013;14:389–394.

38. Wu JW, Hussaini SA, Bastille IM, et al. Neuronal activity enhances tau propagation and tau pathology in vivo. Nat Neurosci 2016;19:1085–1092.

39. Darki F, Peyrard-Janvid M, Matsson H, Kere J, Klingberg T. Three dyslexia susceptibility genes, DYX1C1, DCDC2, and KIAA0319, affect temporo-parietal white matter structure. Biol Psychiatry 2012;72:671–676.

40. Parra Bravo C, Naguib SA, Gan L. Cellular and pathological functions of tau. Nat Rev Mol Cell Biol 2024;25:845–864.

41. Scholl M, Lockhart SN, Schonhaut DR, et al. PET Imaging of Tau Deposition in the Aging Human Brain. Neuron 2016;89:971–982.

